# Consensus guideline for the management of patients with appendiceal tumors: Part 2: Appendiceal tumors with peritoneal involvement

**DOI:** 10.1101/2024.08.30.24309032

**Authors:** PSM Appendiceal Tumor Writing Group, PSM Consortium Group, Kiran K. Turaga

## Abstract

**Background:** Appendiceal tumors comprise a heterogeneous group of tumors which frequently disseminate to the peritoneum. Management of appendiceal tumors is lacking high quality data, given its rarity and heterogeneity. In general, appendiceal tumor treatment is extrapolated in part from colorectal cancer or pooled studies, without definitive evidence of disease-specific benefit. Many practices are controversial and vary widely between institutions. A national consensus update of best management practices for appendiceal malignancies was performed to better standardize care. Herein we present recommendations for management of appendiceal tumors with peritoneal involvement.

**Methods:** As previously described, modified Delphi consensus was performed to update the previous 2018 Chicago consensus guideline. Recommendations were supported using rapid systematic reviews of key issues in surgical and systemic therapy. Key pathology concepts and recommendations were synthesized in collaboration with content experts.

**Results:** A consensus-based pathway was generated for any type of non-neuroendocrine appendiceal tumor with peritoneal involvement. The first round of Delphi consensus included 138 participants and 133 (96%) participated in the second round, with over 90% consensus achieved for all pathway blocks. Key items include recommending evaluation for cytoreduction to most patients with low-grade peritoneal disease who are surgical candidates, and to many patients with high-grade disease, as well as timing of systemic chemotherapy and surveillance protocols. Common pitfalls in pathologic classification and their clinical implications are also presented.

**Conclusion:** These consensus recommendations provide guidance regarding the management of appendiceal tumors with peritoneal involvement, including a review of current evidence in management of recurrent and unresectable disease.

## BACKGROUND

Appendiceal tumors are a diverse group of rare, heterogeneous tumors. Data to guide their management is by nature low in volume and rarely prospective, so expert consensus guidelines remain a necessity to direct care, as was previously done in the 2018 Chicago Consensus.^1–5^ Part 1 of the 2023 consensus guideline update presented management for localized appendix tumors.^6^ This second part presents management of appendix tumors with peritoneal involvement.

Approximately 40-50% of appendiceal tumors present with distant disease at diagnosis, most often mucinous tumor deposits in the peritoneum called pseudomyxoma peritonei (PMP).^7–9^ For any mucinous neoplasms with peritoneal involvement, median overall survival (OS) ranges from 51 to 160 months.^4,10–24^ Moderately- to poorly-differentiated disease has less favorable median OS of 42-66 months, and non-mucinous disease even less, at 18.9-24 months. ^22–29^ Survival of goblet cell adenocarcinoma (GCA) with peritoneal involvement also differs by grade, from a median OS of 98 months for grade 1 and 2 disease, to 33 months for grade 3 disease.^30^

Although estimates vary widely, recurrence is common, with approximately 25% of all comers developing recurrence in the first 1-3 years after surgery.^31–33^ Rates are higher with increasing cellularity and grade of peritoneal disease.^34–38^ In one of the largest studies of recurrence after cytoreduction, Govaerts et al reported disease-free survival plateaus around 6 years after surgery, with 60% of patients with low-grade and 20% of patients with high-grade tumors remaining disease-free.^39^

## METHODS

The methods including the modified Delphi process for the 2023 consensus update of the 2018 Chicago Consensus Guidelines have been previously described in detail.^6,40^

### Consensus Group Structure

The Appendiceal Tumor Working Group included fourteen multidisciplinary disease site experts and a systematic review expert. Two steering committee core members coordinated and revised the pathways (FM, EG). Sixteen trainees (medical students, residents, and fellows) conducted the rapid reviews. Updated pathology recommendations were developed collaboratively with members of the consensus group with gastrointestinal pathology expertise.

### Rapid Review of the Literature

Rapid reviews were conducted of Medline as described in the methods document and the part 1 guideline for management of localized appendiceal tumors.^6,40^ Key question 1 addressed the optimal timing of systemic chemotherapy relative to cytoreduction in non-low grade peritoneal disease of appendiceal origin. The latter two of the three key questions address the following, with detailed search strategies available in supplemental tables 1-2.

1. In patients with unresectable pseudomyxoma peritonei, which management approaches offer the best symptom control and survival benefit (surgical interventions vs. systemic therapy vs. conservative management)? (PROSPERO CRD42023463230)
2. In patients with recurrent peritoneal disease after initial cytoreductive surgery for appendiceal tumors, is repeat cytoreduction with or without hyperthermic intraperitoneal chemotherapy safe and superior to systemic therapy alone or observation? (PROSPERO CRD42023463240)

Reviews were conducted and data extracted according to the published protocols and methods detailed in the Introduction and Methods and part 1 Appendiceal Tumors document, in which key question 1 is also summarized.^6,40^

## RESULTS

### Pathways

Of 138 experts and thought leaders voted on the clinical pathway for appendix tumors with peritoneal involvement, 133 (96%) participated in the second round. The group comprised 96 (70%) surgical oncologists, 20 (14%) medical oncologists, 15 (11%) pathologists, and 7 (5%) experts from other disciplines. The blocks are summarized below with supporting literature incorporated where appropriate.

### Rapid reviews

#### Key question 2: Management of unresectable appendiceal malignancy

For key question 2, 1473 abstracts were screened, 103 included for full-text review, and 15 selected for final inclusion, reporting outcomes of any intervention for the management of initially unresectable pseudomyxoma peritonei of appendiceal origin of any grade. Inclusion and exclusion criteria are detailed in the KQ2 Prisma flow diagram and key (Supplemental Figure 1 and Table 3). Most exclusions were due to overlap with key question 3, recurrent disease. Outcomes were overall and progression-free survival, eligibility for and completeness of cytoreduction, and adverse events. While not amenable to meta-analysis, the included heterogeneous studies explore a range of therapies for unresectable PMP. All but one were retrospective observational studies; the other was a single-arm Phase 2 trial. Six studies reported on systemic chemotherapy,^41–48^ 6 on incomplete cytoreduction or debulking,^46,49–56^ and 7 on intraperitoneal chemotherapy,^45,47,48,50,52,54,56^ with substantial overlap. Results are summarized in Table 1 and quality assessment in Supplemental Table 4.

**Table 1.** Key Question 2: Management Approaches for Initially Unresectable PSM of Appendiceal Origin.

| Author, year, country | Population and inclusion criteria | Tumor grade/type and disease burden | Interventions | Outcomes reported | Adverse events (grade 3/4) |
| --- | --- | --- | --- | --- | --- |
| Studies of exclusively unresectable appendiceal cohorts |  |  |  |  |  |
| **Baron 2022 (USA)**<sup>42</sup> | 72 pts with high grade PMP who underwent aborted/palliative CRS; excluded deaths within 90 days of surgery | All high grade<br><u>SCT:</u><br>8/20 with SRCs<br>1/20 goblet cell<br><u>No SCT:</u><br>29/50 with SRCs<br>8/50 goblet cell | **SCT:**<br>20/72 with median 12 cycles of 5-FU/capecitabine ± oxaliplatin ± irinotecan<br>10/20 with bevacizumab<br>**No SCT:**<br>52/72 | **<u>OS, Repeat CRS</u>**<br>- Median OS 26 mo for SCT vs 12 mo for no SCT (HR 0.22, 95% CI 0.08-0.66; p=0.007)<br>- 2/20 SCT group with reattempted CRS, 1/20 completed CRS | Grade 3-4 complications 3/20 SCT vs. 3/52 no SCT |
| **Choe 2015 (US)**<sup>43</sup> | 130 pts treatment naive to SCT and not amenable to complete CRS due to extensive disease, progression after prior CRS, or assessed low benefit from surgery | 37/130 well-diff<br>43/130 mod-diff<br>50/130 poor-diff<br>33/130 with signet ring cells | **SCT with anti-VEGF or anti-EFGR:**<br>59/130 total with bevacizumab, all 5-FU based<br>5/130 with cetuximab, also 5-FU based<br>1/130 with panitumumab<br>**Standard SCT without biologic:**<br>51/130 standard 5-FU or Cape based<br>14/130 other | **<u>OS, PFS, Response</u>**<br>- Median PFS 9 mo with bevacizumab vs 4 mo without biologic (HR 0.69, 95% CI 0.46-0.995; p = 0.047)<br>- Median OS improved by 34 mo with bevacizumab vs without biologic (HR 0.49, 95 % CI 0.25–0.94; p = 0.03)<br>- Anti-EGFR associated with reduced median OS by 18-20 mo<br>- Response to SCT (imaging/clinical): 87% with bevacizumab vs 60% without biologic | NR |
| **Delhorme 2016 (France)**<sup>49</sup> | 39 pts with unresectable disease | 13/39 DPAM<br>12/39 PMCA-I<br>14/39 PMCA | **Debulking (Incomplete CRS):**<br>23/39 underwent >80% (major) debulking<br>24/39 w/prior CRS<br>17/39 w/prior SCT | **<u>OS, PFS, Symptom resolution</u>**<br>- Median OS 55.5 mo; 5-yr OS 46%<br>- Median PFS 20 mo; 5-yr PFS 11%<br>- Resolution of PMP-related symptoms in 14/39 pts | NR |
| **Farquharson 2008\* (UK)**<sup>44</sup> | Phase 2 experimental trial 40 pts with unresectable PMP, adequate performance status, and life expectancy > 3 mo | 27/40 DPAM<br>10/40 PMCA-I/-D<br>3/40 PMCA | **SCT:** 8 cycles every three weeks of:<br>MMC 7 mg/m<sup>2</sup> IV on day 1 and capecitabine 1250 mg/m<sup>2</sup> BID on days 1-14 | **<u>OS, Attempt at CRS, and Response</u>**<br>- 1-yr OS: 84%; 2-yr OS: 61%<br>- Attempt at CRS: 2/40<br>- Response to SCT (imaging): 15/40 (38%) | Grade 3 events in 12/277 cycles, grade 4 events in 4/277 cycles |
| **Mangieri 2022 (USA)**<sup>50</sup> | 93 pts with incomplete CRS (R2b or R2c) | 39/93 low grade<br>48/93 high grade<br>6/93 NOS | **CRS-HIPEC:** 43/93 (mitomycin C or oxaliplatin)<br>**CRS alone:** 50/93 | **<u>OS and PFS (CRS-HIPEC vs. CRS alone)</u>**<br>**Median OS**<br>- All: 2.3 yrs vs 1.2 yrs (p=0.016)<br>- Low-grade: 1.9 yrs vs 1.2 yrs (p=0.004)<br>- High-grade: 1.15 yrs vs 1.6 yrs (p=0.484)<br>**Median PFS**<br>- All: 1.3 yrs vs 0.6 yrs (p<0.0001)<br>- Low-grade: 1.80 yrs 0.63 yrs (p=0.004)<br>- High-grade: 1.19 yrs 0.43 yrs (p=0.016) | NR |
| **Murphy 2007 (UK)**<sup>51</sup> | 40 pts not amenable to complete CRS | 32/40 low grade<br>8/40 adenocarcinoma | **Open-and-close:** 6<br>**Major debulking:** 34 CC-2/3 | **<u>Mortality and OS</u>**<br>**Open-and-close:** 2/6 30-day mortality; OS < 6 mo<br>**Major debulking:** 1/34 30-day mortality; 2-yr OS 57% | NR for unresectable group |
| **Shapiro 2010 (USA)**<sup>45</sup> | 54 pts not amenable to complete CRS due to extensive disease, progression after prior CRS, or assessed low benefit from surgery | 24/54 well-diff<br>11/54 mod-diff<br>15/54 mod-poor diff<br>4/54 undetermined | **SCT:**<br>38/54 received 2 cycles; most 5-FU or capecitabine based +/- platinum; some irinotecan regimens (7/54) and 5/54 gefitinib; 11/54 received biologic therapy<br>**SCT and IPCT:** | **<u>OS and Response</u>**<br>- Median OS 40 mo for SCT vs. not reached at 80 mo for SCT + IPCT<br>- 3-yr OS 50.6% for SCT vs. 73.4% for SCT + IPCT (p=0.0495)<br>- Response to SCT (imaging/clinical): 56% (No difference between groups) | NR |
|  |  | 18/54 (33%) with signet ring cells | 16/54 also received IP mitomycin C |  |  |
| **Sideris 2009 (Canada)**<sup>46</sup> | 37 pts undergoing CRS + IPCT with curative intent, of which 12 pts deemed initially unresectable | NR for pts with unresectable disease | **SCT:** (6 mo FOLFOX/FOLFIRI) followed by CRS + HIPEC: 2/12 pts<br>**Palliative surgery and SCT NOS:** 10/12 pts | **<u>Attempt at CRS</u>**<br>Both patients who received 6 mo SCT underwent complete CRS + HIPEC; survival data not specified for unresectable disease | NR |
| **Smeenk 2007 (Netherlands)**<sup>52</sup> | 10 pts undergoing CRS +/- IPCT for disease recurrence or progression after prior CRS | 7/10 DPAM<br>3/10 PMCA-I | **Incomplete cytoreduction + HIPEC:**<br>1/10 later had systemic chemotherapy for recurrence; further results not specified | **<u>Progression</u>**<br>- 7/10 progressed - 5/7 DPAM and 2/7 PMCA<br>- Median PFS 6-9 mo based on extrapolated survival curve | NR |
| **Sugarbaker 2022 (USA)**<sup>53</sup> | 264 pts with PMP and incomplete CRS | 83/264 low-intermediate grade (LAMN/MACA-I)<br>85/264 grade 1-2 adenocarcinoma<br>73/264 grade 3 or with SRCs | **Incomplete CRS (CC2-3)**<br>SCT before CRS: 107/264<br>HIPEC: 14/264<br>EPIC: 147/264 | **OS**<br>- Median OS 1.8 yrs for whole cohort<br>- Median OS 1.5 yrs for SCT before CRS vs. 2.0 yrs for no SCT before CRS (HR 1.4, 95% CI 1.1-1.8; p = 0.008) | 2 post-procedural deaths in HIPEC+EPIC<br>26/264 with grade 4 complications |
| **Trilling 2021 (Canada)**<sup>54</sup> | 8 pts with low-grade PMP and predicted complete CRS after two sequential surgeries | 4/8 DPAM<br>4/8 PMCA-I | **Two-step CRS-HIPEC**<br><u>1<sup>st</sup> CRS:</u> 8/8 CC-2<br><u>2<sup>nd</sup> CRS:</u> 5/8 CC-0; 3/8 CC-1; 1/8 unresectable | **<u>OS</u>**<br>- Median OS not reached at median follow-up of 53.8 mo<br>- 3-yr OS 100%, 5-yr OS 85.7%<br>- 1 pt lost to follow-up | NR |
| **Vierra 2022 (USA)**<sup>47</sup> | 13 pts with PMP and ≥ 1 incomplete CRS ± SCT or HIPEC, receiving celecoxib-myrto combination | 9/13 low-grade<br>4/13 high-grade | **Celecoxib-myrto**<br>- 11/13 HIPEC<br>- 7/13 with multiple previous cycles of SCT | **<u>OS, PFS, Response</u>**<br>- Median OS 27 mo<br>- Median PFS 16 mo<br>- Tumor marker response - 9/13 | Mortality 2/13 in 8 mo; Bowel obstructions 3/13 |
| Studies including unresectable appendiceal cohorts as a subpopulation, compared to resectable |  |  |  |  |  |
| **Berger 2021 (USA)**<sup>48</sup> | 7 pts who underwent iterative HIPEC for high-grade appendiceal ex-goblet cell adenocarcinoma with unresectable peritoneal disease; s/p 3-6 months of SCT without progression | 4/7 Tang B AEGA<br>3/7 Tang C AEGA | **Iterative HIPEC:** Open/laparoscopic mitomycin C HIPEC, 2-3 cycles per pt<br>Outcomes compared with institutional control groups of pts with high grade appendiceal tumors who underwent<br>- SCT only (n = 16)<br>- Complete CRS-HIPEC (n = 7) | **<u>OS, CRS-HIPEC reattempt</u>**<br>- Median OS 24.6 mo for HIPEC vs 7.9 mo for SCT only (p = 0.005) vs 16.5 mo for CRS-HIPEC (p = 0.62)<br>- 2/7 reattempted CRS-HIPEC, not complete<br>- No significant change in PCI across HIPEC intervals | IHIPEC: 0 procedural complications, 5 90-day readmissions across 4 patients |
| **Dayal 2013 (UK)**<sup>55</sup> | 205 pts who underwent maximal tumor debulking | 61/205 high grade<br>3/205 with signet ring cells | **Maximal Tumor Debulking** | **<u>OS</u>**<br>Median OS 32.8 mo; 3-, 5-, and 10-yr OS 47%, 30%, and 22% respectively | 167 complications<br>22/205 return to OR |
| **Polanco 2016 (US)**<sup>56</sup> | 97 pts with high grade disease, of which 21 pts underwent incomplete CRS (CC2-3); compared outcomes in pts with high volume vs. low volume disease | All high grade<br>32/97 had signet ring cells | **CRS ± HIPEC:**<br><u>High volume disease</u> (SPCI ≥ 12):<br>15/54 CC-2/3<br>43/54 HIPEC<br>32/54 prior SCT; 17/54 adjuvant SCT<br><u>Low volume disease</u> (SPCI < 12):<br>6/43 CC-2/3<br>42/43 HIPEC<br>31/43 prior SCT; 8/24 adjuvant SCT | **<u>OS and PFS (not specified for CC2-3)</u>**<br>- Median OS 17.42 mo in high vs 42.4 mo for low volume (p=0.009)<br>- Median PFS 9.6 mo for high vs 14.2 mo for low volume (p=0.002)<br>- Comparable median OS (56 mo vs. 52 mo, p=0.73) and PFS (20 mo vs. 19 mo, p = 0.39) between high vs. low volume when CC0 achieved | NR |

\*Except for the indicated study, all studies were retrospective cohort studies of prospectively collected data, whether in a prospective database or of previously recorded patient chart information.

Better outcomes were generally reported across the included studies for patients who underwent cytoreductive surgery or major tumor debulking compared to those who did not, but all studies included here are observational and retrospective.^49,51,53,55,56^ Patients with sufficient physiologic reserve to undergo invasive procedures, disease biology that is sufficiently indolent to allow them to maintain surgical candidacy after courses of systemic therapy, and disease burden and distribution that does not initially preclude attempted resection all contribute independently to better survival outcomes, making it impossible to account for the true impact of selection bias.

For cohorts including low-grade disease, six studies demonstrated either increased survival compared to observation or an acceptable survival and morbidity profile for tumor debulking or incomplete cytoreduction.^49–51,53–55^ Intraperitoneal chemotherapy was part of the treatment regimen in five retrospective, non-randomized studies involving low-grade disease, two of which demonstrated a survival advantage of regimens with intraperitoneal chemotherapy over regimens without.^45,47,50,52,54^

For patients with high-grade disease, four included observational studies report either improved survival or eligibility for cytoreduction after upfront systemic therapy, although two studies do not explicitly report which outcomes were associated with high- vs low-grade; most strikingly, Baron et al reported median OS of 26 months with systemic chemotherapy SCT compared to 12 months without.^42–44,46^ While Sugarbaker et al reported longer survival among patients undergoing upfront incomplete cytoreduction instead of SCT, as an observational study over 22 years, it is likely that patients with more indolent tumor biology underwent upfront surgery and that patients undergoing upfront SCT received regimens that would now be considered obsolete.^53^ Three studies reported a survival benefit associated with IPCT, and another reported increased survival in patients undergoing iterative HIPEC (iHIPEC) in high-grade goblet cell disease.^45,48,50^ As the authors reporting on iHIPEC themselves discuss, the likely impact of selection bias, both in terms of inclusion only of patients able to undergo multiple surgical procedures but also the exclusion of individuals whose disease progressed during SCT, it is difficult to make generalizations about the benefit of iHIPEC, but it is worth noting that survival in their experimental cohort did exceed historical comparators.^48^

Another small study evaluated celecoxib-myrtol combination therapy along with IPCT for disease of any grade, with modest biochemical response in 9 of 13 patients.^47^ Bevacizumab has been associated with favorable progression-free and overall survival for any grade in one included study and another more recent study by Hornstein et al, described in part 1.^43,57^ A recent prospective, randomized trial with a crossover design, discussed in part 1, demonstrated that systemic chemotherapy with 5FU-based regimens was no better than observation in terms of survival for individuals presenting with unresectable low-grade mucinous disease.^3^ While excluded from the review because most participants had recurrent, not initially unresectable disease, it supports our consensus that conventional cytotoxic chemotherapy regimens are unlikely to provide benefit for most patients with unresectable low-grade peritoneal disease.

#### Key Question 3: Repeat Cytoreduction for Recurrent Appendiceal Malignancy

For key question 3, 191 abstracts were screened, 36 included for full-text review, and 9 selected for final inclusion, reporting outcomes of repeat cytoreduction for recurrent pseudomyxoma peritonei of appendiceal origin of any grade after previous cytoreduction. Inclusion and exclusion criteria are detailed in the KQ3 Prisma flow diagram (Supplemental Figure 2 and key in Table 5). Most exclusions were due to reporting outcomes collectively with non-appendiceal disease, or overlap with other studies or key question 2, which evaluated initially unresectable rather than recurrent disease. Meta-analysis was not possible, but summation of outcomes including overall survival and adverse events support an overall survival advantage of repeat cytoreductive surgery for carefully selected patients with appendiceal tumors experiencing relapse after initial CRS, compared to non-surgical management.^31,58–65^ Results are summarized in Table 2 and quality assessment in Supplemental Table 6. All were retrospective observational studies.

**Table 2.** Key Question 3: Repeat cytoreductive surgery in the management of recurrent peritoneal disease.

| Author, year, country | Population: Tumor/PMP grade | Intervention (Sample size) |  | Outcomes |  |  |  |  |
| --- | --- | --- | --- | --- | --- | --- | --- | --- |
|  |  |  |  | Median OS (months) |  |  | OS HR (95%CI) | Grade 3-4 adverse events after repeat CRS |
|  |  | Repeat CRS | Non-surgical | Repeat CRS | Non-surgical | Log rank p-value |  |  |
| Ahmadi 2021<sup>¶</sup> (UK)<sup>59</sup> | low-grade, high-grade, and adenocarcinoma | 145 | 119 | - | - | - | 0.41 (0.23-0.74) | 14.0% of 114 (41 unknown) |
| Delhorme 2017 (France)<sup>64</sup> | DPAM, PMCA-I, PMCA, and unknown | 47 | 19 | Not reached | ~18* | <0.05 | - | 37.9% |
| Karpes 2020<sup>†</sup> (Australia)<sup>61</sup> | DPAM | 44 | 176 | 125.5 | 248.7 | 0.15 | 1.11 (0.54-2.29) | 44.1% |
|  | PMCA | 58 | 184 | 90.7 | 55.6 | 0.03 | 0.57 (0.33-0.95) |  |
| Lopez-Ramirez 2022<sup>‡</sup> (USA)<sup>62</sup> | Overall | 55 | 55 | 80.2 | 36.2 | <0.001 | - | 27.3% |
|  | LGMCP | 36 | 36 | 174.1 | 51.9 | <0.001 | 0.32 (0.16-0.63) | - |
|  | HGMCP | 13 | 13 | 42.0 | 12.4 | 0.02 | 0.36 (0.13-0.98) | - |
|  | HGMCP-S | 6 | 6 | 15.4 | 8.1 | 0.61 | 0.84 (0.25-2.79) | - |
| Valenzuela 2022<sup>§</sup> (USA)<sup>58</sup> | LAMN | 58 | 85 | 227.1 | 54.5 | <0.001 | - | - |
| Studies including participants with recurrent appendiceal disease as a sub-cohort, also including progressive disease and/or non-appendiceal cohorts |  |  |  |  |  |  |  |  |
| Kitai 2020 (Japan)<sup>63</sup> | AM, DPAM, PMCA, PMCA-S | 10 | 3 | Not reached | ~28* | <0.05 | - | - |
| Kong 2021<sup>#</sup> (Australia)<sup>31</sup> | AM | 2 | 0 | 64.7 | 38.7 | <0.001 | - | 30% complications (not graded) |
|  | Low grade | 24 | 21 |  |  |  |  |  |
|  | High grade | 4 | 9 |  |  |  |  |  |
| Powers 2020 (USA)<sup>60</sup> | 78.5% appendiceal, not otherwise specified | 126 | 243 | 73 | 36 | 0.001 | - | 30.7% |
| Yan 2007 (USA)<sup>65</sup> | DPAM, PMCA-I, PMCA | 98 | 13 | Not reached | ~34* | <0.001 | - | - |
| * Median OS extrapolated from Kaplan-Meier curve<br>† Patients who underwent repeat CRS were younger and underwent fewer complete cytoreductions<br>‡ Propensity score matched institutional controls for non-surgical management<br>§ Patients who underwent repeat CRS were younger, had better performance status, underwent more complete initial CRS, and had a longer time to recurrence from initial CRS<br>¶ Patients with high tumor grade, early recurrence, and raised tumor markers at recurrence were less likely to have repeat surgery; Potential overlap between intervention groups not otherwise specified or explained by study authors<br># Patients offered repeat CRS were younger and had more low grade disease<br>All studies were retrospective cohort studies of prospectively collected data, whether in a prospective database or of previously recorded patient chart information.<br>**Abbreviations:** PMP: pseudomyxoma peritonei; OS: overall survival; CRS: cytoreductive surgery; LAMN: low-grade appendiceal mucinous neoplasm; DPAM: disseminated peritoneal adenomucinosis; PMCA: peritoneal mucinous carcinoma peritonei; LGMCP: low-grade mucinous carcinoma peritonei; HGMCP: high-grade mucinous carcinoma peritonei; -S: with signet ring cells; AM: acellular mucin; -I: intermediate grade, referring to Ronnett classification; HR: hazard ratio; OS: overall survival |  |  |  |  |  |  |  |  |

Notably, two high-quality comparative studies provided stratified results based on tumor grade. Karpes et al. demonstrated improved overall survival in multivariable analysis in patients undergoing repeat CRS versus non-surgical management for PMCA (high-grade disease) (HR 0.57) but not for DPAM (low-grade or acellular disease).^61^ Lopez-Ramirez et al. compared outcomes between patients who underwent repeat CRS and a propensity-score matched cohort of patients who did not, and noted a survival benefit of repeat CRS for both low- and high-grade disease (HR 0.32 and 0.36, respectively), excluding those with signet ring cells.^62^ The incidence of grade 3-4 postoperative adverse events following repeat CRS ranged from 14.0% to 44.1% across studies.^59,61,62^

It is difficult to separate selection and publication biases from the necessarily careful selection of candidates for repeat CRS. Collectively, the included studies suggest benefit from repeat CRS in terms of overall survival, and also describe patient selection for repeat CRS as those with more favorable patient and disease characteristics including younger age, better performance status, lower-grade disease, longer intervals to disease recurrence, and less frequent elevation of tumor markers at recurrence.^31,58,59,61^ This emphasizing the need for individualized decision-making regarding repeat CRS by weighing the risks and benefits of major surgery carefully.

## PATHOLOGIC CLASSIFICATION OF APPENDICEAL TUMORS

The pathologic classification and grade of appendix tumors remains a major determinant of survival, in the context of growing but still incomplete understanding of molecular tumor biology.^27,66–70^ Consequently, our consensus pathway recommendations are stratified by tumor pathology (Table 4). The historical evolution of pathologic classification systems informs understanding of the crosstalk between them and the key issues that clinicians may encounter with interpreting pathologic designations. New developments in molecular tumor biology and genomics suggest future develops in prognostic stratification and identification of treatment targets.

### Historical perspective

Prior to the 1970s, the term “mucocele” was frequently used to describe dilated appendices filled with mucin.^71–75^ When a mucocele perforated and was associated with mucin extrusion, if the mucin continued to accumulate after appendectomy, it was considered a “malignant mucocele,” which was often described as being lined by villiform or undulating mucinous epithelium with elongated pseudostratified atypical nuclei.^76^ Over the next several decades, the terminology for colonic adenomas was extrapolated to the appendix, and the concept of an appendiceal “adenoma,” or “cystadenoma” if cystically dilated, became entrenched in appendiceal tumor nomenclature, although it generated controversy as to the benign character of the lesions when they ruptured and disseminated as pseudomyxoma peritonei.^77,78^ Adding to the controversy was the fact that the pseudomyxoma peritonei resulting from these low-grade tumors is histologically bland, unlike most peritoneal carcinomatosis, leading to considerable debate over whether classic low-grade pseudomyxoma peritonei represents a form of peritoneal carcinomatosis or a separate benign, albeit progressive, intra-abdominal process. Biologically, classic PMP and carcinomatosis have different disease distribution, outcome, and tempo of progression: low-grade PMP coats the surfaces of organs, follows a pattern of peritoneal distribution known as the “redistribution phenomenon”, and is slowly progressive, whereas peritoneal carcinomatosis invades organs and is rapidly fatal.^17^

Some pathologists accepted that cystadenomas might rupture and result in peritoneal implants of benign neoplastic epithelium, and therefore classified the appendiceal lesion as ruptured adenomas and the peritoneal tumor as “disseminated peritoneal adenomucinosis,” implying that the peritoneal tumors were benign.^17^ Other pathologists objected to using any benign terminology.^79,80^ They argued that neoplastic epithelium growing within the peritoneal cavity was malignant and therefore a form of carcinomatosis, regardless of its bland microscopic appearance, and therefore that the appendiceal tumor that produced it must be malignant, even if a pathologist cannot recognize it as such.^78,81,82^. This viewpoint classified any appendiceal tumor associated with PMP as adenocarcinoma, even if it lacked features of invasive adenocarcinomas that arise elsewhere in the gastrointestinal tract. In 1995, Carr and Sobin published a seminal work in which they classified appendiceal mucinous tumors as mucinous adenomas if confined to the mucosa, and as adenocarcinoma if associated with any growth of viable cells outside the appendix. Tumors that pushed into the underlying appendix wall or acellular mucin pools on the serosa but were not associated with PMP were classified as mucinous tumors of uncertain malignant potential (UMP), to acknowledge the difficulty in identifying invasion.^81^

In 2003, Misdraji and colleagues coined the term “low-grade appendiceal mucinous neoplasm” (LAMN) to describe the full spectrum of these tumors, arguing that the tumors classified variously as cystadenoma, mucinous neoplasm of uncertain malignant potential, and occasionally adenocarcinoma were histologically indistinguishable, with or without peritoneal spread.^83^ Using this terminology, LAMNs can be staged just as any tumor, and the biologic potential of a given tumor can be predicted based on its stage. Subsequently, the term high-grade appendiceal mucinous neoplasm (HAMN) was introduced for tumors with pushing invasion but with high-grade cytology.^84^ LAMN and HAMN are now recognized as the tumors most often responsible for pseudomyxoma peritonei, and the presence of PMP no longer mandates classification as adenocarcinoma. Rather, to qualify as adenocarcinoma, a tumor must demonstrate infiltrative-type invasion typical of adenocarcinomas elsewhere in the gastrointestinal tract.

Commensurate with changes in the nomenclature of appendiceal primary tumors, the terminology and grading for PMP went through various changes. Historically, the term pseudomyxoma peritonei was reserved for the clinical syndrome of progressive mucinous ascites, and was most often applied to peritoneal tumors with abundant mucin, dissecting fibrosis, and occasional neoplastic but cytologically bland mucinous epithelial cells.^85^ Ronnett and co-investigators coined the term “disseminated peritoneal adenomucinosis” for low-grade PMP, a term that suggests that low-grade PMP is benign mucinous neoplasia; distinct from high-grade peritoneal mucinous carcinoma (PMCA) which had a worse prognosis.^17^ Their initial paper also had a group of tumors that did not fit neatly into this framework, and they called this group peritoneal mucinous carcinomatosis with intermediate or discordant features (PMCA-I/D). This heterogeneous group included tumors with features intermediate between DPAM and PMCA, but also tumors in which the grade of the appendiceal primary and the peritoneal tumor were discordant.^17^ Some pathologists did not accept DPAM for low-grade PMP, and reported all PMP using malignant terms, such as “mucinous carcinoma peritonei (MCP), low-grade” or “MCP high-grade.”^85^ Over the ensuing years, studies demonstrated that the presence of signet ring cells in PMP conferred a worse prognosis, and this warranted a three-tier grading system, with grade 3 reserved for tumors with signet ring cells.^14^ In the past few years, the World Health Organization, in an attempt to introduce terminology that would be acceptable to all pathologists, adopted the term pseudomyxoma peritonei for all peritoneal mucinous tumors, regardless of grade, in order to avoid confusion as to whether PMP is a benign or malignant condition.^86^ Mucinous carcinoma peritonei was an acceptable alternative.

Finally, another tumor that underwent nomenclature changes in the last two decades is goblet cell adenocarcinoma. This unique tumor is composed of tubules of goblet-like cells, enterocytes, and occasional Paneth-like cells. These tumors may show scattered endocrine cells by immunohistochemistry, and are considered to be amphicrine tumors, showing both endocrine and exocrine differentiation.^87^ Although classic goblet cell tumors are low grade, they can dedifferentiate, and most often these high-grade tumors resemble poorly differentiated adenocarcinomas or signet ring cell carcinoma.^88^ Prior to 1990, the full spectrum of tumor grades was classified as “goblet cell carcinoid”, resulting in their behavior being poorly predicted by their nomenclature.^89^ The labeling of these tumors as “carcinoid” was a reflection of their bland appearance and their ability to infiltrate the appendix circumferentially without forming a mass lesion. In 1990, Burke and others at the Armed Forces Institute of Pathology published the first grading system for these tumors.^88^ Tumors with less than 25% “carcinomatous” growth were biologically indolent and were classified as “goblet cell carcinoid” whereas those with greater than 50% carcinomatous growth were classified as mixed adenocarcinoma-carcinoid. In 2008, Tang and coinvestigators proposed a different grading system for these tumors. Low-grade tumors were classified as “goblet cell carcinoid”. High-grade tumors were classified as “adenocarcinoma ex goblet cell carcinoid”, with two subtypes.^90^ Although this classification system became well known, it was challenging to apply in practice due to the heterogeneous nature of goblet cell tumors and the somewhat vague criteria for distinguishing the various grades. Unfortunately, the term “adenocarcinoma ex-goblet cell carcinoid” gave the impression that, in these high-grade tumors, an adenocarcinoma is arising from an endocrine tumor, which is not the case. There also continued to be confusion as to whether neuroendocrine staging systems or therapies should be applied to these tumors. Yozu and colleagues reclassified these tumors as “goblet cell adenocarcinoma” in 2018, in recognition of their being more closely aligned with adenocarcinoma than with neuroendocrine tumors.^91^ They also proposed a grading system for these tumors that depended on the extent to which the tumor recapitulated the low-grade tubular morphology. In the most recent WHO classification, this nomenclature and grading system was adopted for this unique appendiceal tumor.^92^

### Interpreting the crosstalk between classification systems

Although the WHO 2019 classification (5^th^ edition) should be used as the mainstay of appendiceal tumor classification to facilitate uniform clinical management and investigation, prior terminology will still be encountered due to clinician familiarity, the large body of literature using older terminology, and patients carrying these diagnoses. It is therefore necessary for clinicians to understand the crosstalk between classification schemes to appropriately stratify patients to risk and treatment groups. Expert interpretation of this crosstalk is presented in Table 3, although perfect alignment between classification schemes is not feasible due to different histologic criteria, weighting, and thresholds.^14,35,83,84,86,92–96^

**Table 3.**
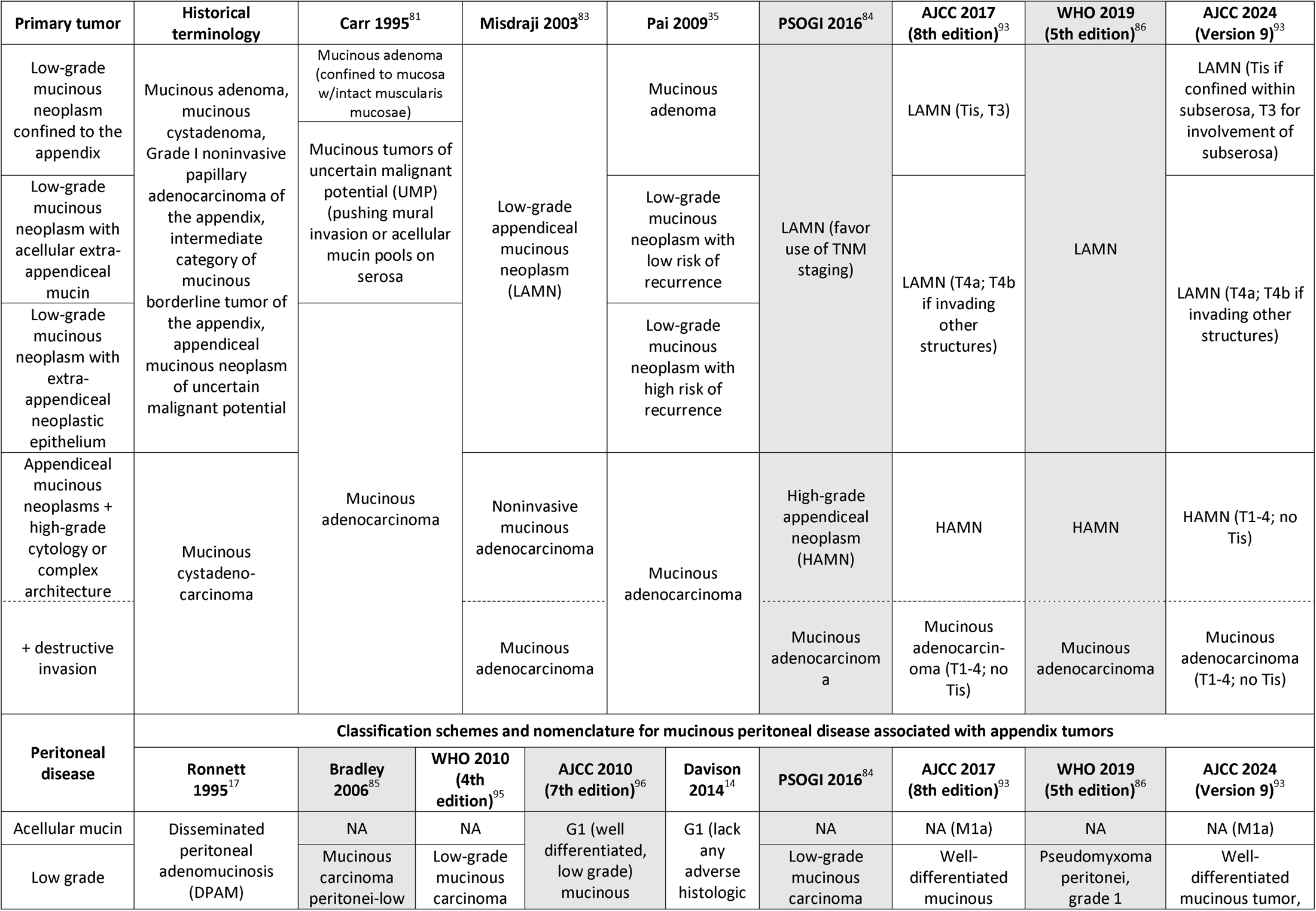
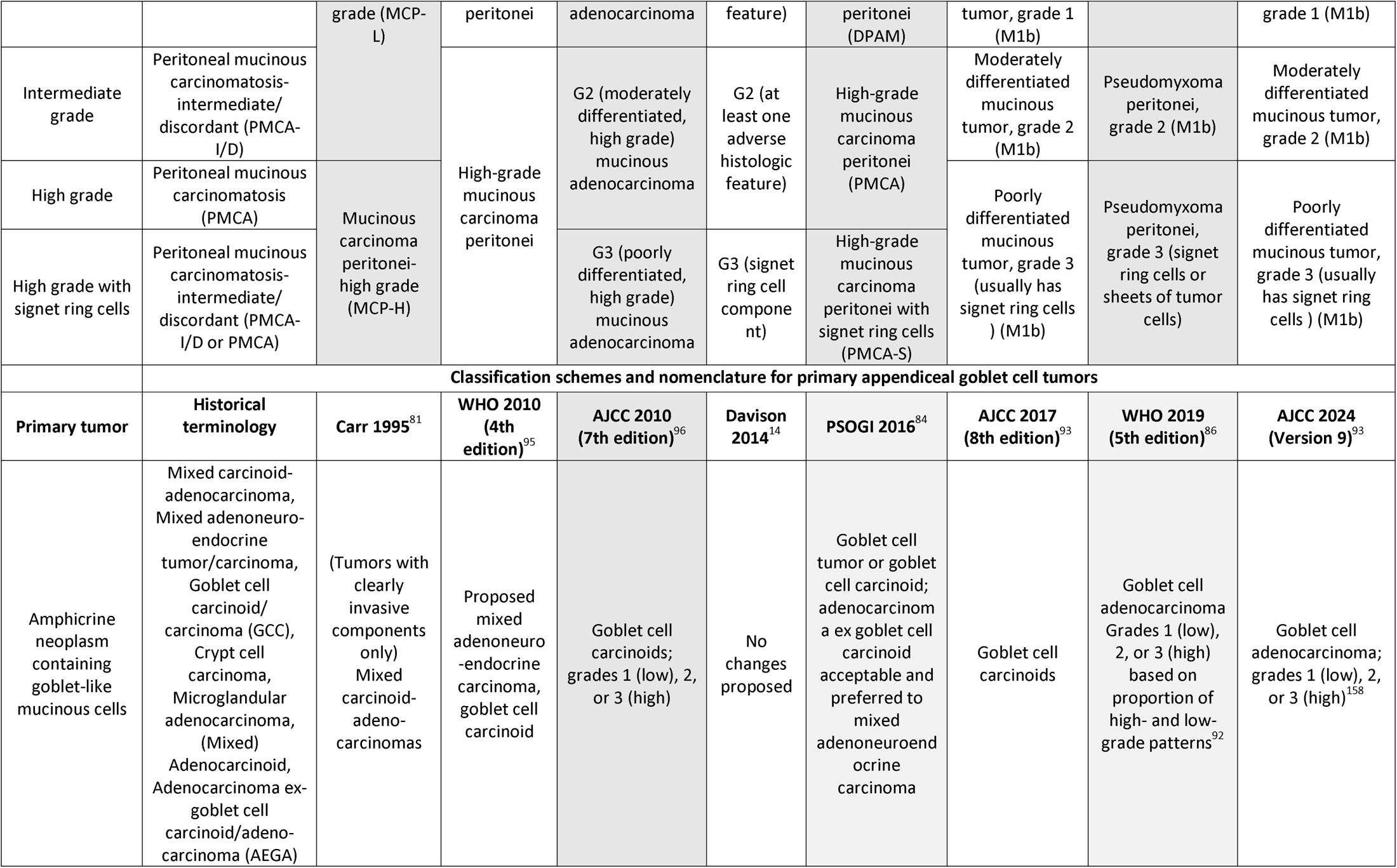
Crosstalk of classification terminology for appendix tumors.

Low-grade appendiceal mucinous neoplasm is the WHO term for a low-grade mucinous tumor that shows pushing type invasion.^97^ Historical diagnoses including appendiceal mucocele, adenoma, or cystadenoma, listed in the leftmost column of Table 3, likely indicate LAMN by today’s standards, particularly a diagnosis of ruptured appendiceal adenoma. High-grade appendiceal mucinous neoplasm or HAMN is the current terminology for what was often described as non-invasive adenocarcinoma or cystadenocarcinoma in prior publications, including the article by Misdraji and others that introduced the term LAMN.^35,83,84,98^

By today’s standards, a diagnosis of adenocarcinoma in the appendix requires at least focal infiltrative invasion. However, the diagnosis of adenocarcinoma has been used for any tumor associated with pseudomyxoma peritonei, as codified in the 1995 Carr paper that proposed that any appendiceal tumor with unequivocal growth of neoplastic cells in the peritoneal cavity be classified as adenocarcinoma.^81^ Today, most tumors that are associated with the pseudomyxoma peritonei are either LAMN or HAMN, but this may be confusing to some pathologists, who may incorrectly diagnose tumors with pseudomyxoma peritonei as adenocarcinoma. In other situations, a pathologist may interpret “pushing invasion” with artifact introduced by plane of section as infiltrative invasion, resulting in a diagnosis of adenocarcinoma. Alternatively, they might interpret localized pseudomyxoma peritonei on the appendiceal serosa as mucinous adenocarcinoma, resulting in misdiagnosis of adenocarcinoma; in the current WHO classification, a diagnosis of adenocarcinoma requires infiltrative type invasion in the appendix itself.^92^ Table 3 shows where each classification system delineates adenocarcinoma.

Goblet cell adenocarcinoma (GCA) is the current terminology for all three grades of goblet cell tumor. In the paper by Burke et al, goblet cell carcinoid corresponds to GCA grade 1 whereas mixed adenocarcinoma-carcinoid is equivalent to GCA grade 3.^88^ The classification by Tang and colleagues is more difficult to translate into WHO terminology.^90^ Their goblet cell carcinoid is roughly equivalent to GCA grade 1. Adenocarcinoma ex-goblet cell carcinoid, signet ring cell type in the Tang classification might correspond to GCA grade 1 or 2, depending on the extent to which the tumor is disorganized, whereas adenocarcinoma ex-goblet cell carcinoid, poorly differentiated carcinoma type is likely to translate to GCA grade 3 but can fall anywhere on the range from GCA grade 1 to 3 depending on the extent of poorly differentiated carcinoma. Table 3 does not expound upon the specific grades, as our treatment recommendations do not differ between grades, but also lists historical terminology used for GCA.

Pseudomyxoma peritonei (PMP) grade 1 corresponds to disseminated peritoneal adenomucinosis (DPAM) in the Ronnett classification, and grade 2 or 3 corresponds to their peritoneal mucinous carcinoma (PMCA).^17^ Their intermediate category (PMCA-I) likely corresponds to PMP grade 1 to focal 2. In the Peritoneal Surface Oncology Group International system, low grade mucinous carcinoma peritonei is equivalent to PMP grade 1, high-grade mucinous carcinoma peritonei is equivalent to PMP grade 2, and high-grade mucinous carcinoma peritonei with signet ring cells is equivalent to PMP grade 3.^84^ The PMP portion of table 3 illustrates these interactions by classification scheme.

### Areas of concern for the clinician

Clinicians caring for patients with appendix tumors should be aware that the pathologic evaluation of these tumors is challenging. Studies of patients referred to high-volume centers for oncologic care demonstrated 24.8-27.8% discordance in diagnosis between referring institutions and high-volume centers, and nearly 50% discordance in grading, with up to 30% discordance associated with change of clinical management.^99–101^ Review by an expert GI pathologist should be considered, particularly if certain common pitfalls may have affected the patient’s initial evaluation.

Perhaps the most common error that affects patient management is mistaking post-inflammatory mucosal hyperplasia or diverticular disease in the appendix as low-grade appendiceal mucinous neoplasm.^102^ If a patient is diagnosed with LAMN that is reported as perforated, but does not have widespread pseudomyxoma peritonei, expert review is suggested, as many of these cases may in fact be benign mimics of LAMN. Similarly, if the pathologist reports two tumors in the appendix, such as a neuroendocrine tumor together with LAMN, they may be over-interpreting obstructive hyperplastic changes in the appendiceal mucosa as LAMN, and expert review is justified.

Unusual pathologies also merit consideration of expert review. For example, LAMN arising from a serrated lesion is a controversial entity, and expert review may clarify the diagnosis.^103^ Also, bona fide cases of high-grade appendiceal mucinous neoplasm are uncommon.^84,104^ If the clinical management of the patient depends on whether the tumor is LAMN or HAMN, expert review is suggested. Non-mucinous adenocarcinomas may be difficult to differentiate on a purely histologic basis from cecal and other right colon cancers, particularly in patients with obliterated or obscured anatomy, but have significantly different biology. ^105–107^ Specifically, signet ring cell and high-grade primary goblet cell adenocarcinoma have similar appearances and may also be mistaken for metastatic disease from other primary sites, and the clinician should have a low threshold for requesting additional pathology review. ^108,109^

In terms of peritoneal disease, the location and extent of extra-appendiceal mucin and neoplastic epithelium should be clearly reported in order to ensure the correct management algorithm is followed. In this consensus guideline, where recommendations refer specifically to localized acellular mucin, we recommend clinicians use a definition that involves disease only as distant as the meso-appendiceal fold and peri-appendiceal recesses, and we recommend referring to the peritoneal disease pathway for any cellular or more widespread mucinous disease.^6^ Difficulty in making this determination may arise on the surgeon’s side because of inflammation, obscured anatomy, and other intraoperative obstacles, but may also be due to challenges with pathologic evaluation; in the latter case, secondary review may be helpful. The presence of signet ring cells is also fraught, as “pseudo” signet ring cells may be found in low-grade PMP. Based on limited data, signet ring cells floating in mucin pools may be less biologically important than signet ring cells infiltrating tissue, and if this is not clear in the pathologic report, clarification or expert review should be requested.^110^ Finally, the grade of the appendiceal primary and the peritoneal tumor is usually concordant, but if it is reported as discordant (such as a high-grade appendiceal tumor with low-grade PMP or vice versa), expert review is prudent.^111^ Prognosis and management recommendations follow the grade of the peritoneal disease.^69,112^

An ongoing issue in the evaluation of PMP is the assessment of disease progression and treatment effect, which impacts the evaluation of patients for surgery. The peritoneal regression grading score (PRGS) was proposed to address this issue using an aggregate measure of intraoperative macroscopic findings and histologic features from biopsies in each of four quadrants.^113^ The utility and generalizability of the PRGS is still not entirely clear, but at least one validation study demonstrated that in combination with peritoneal cytology, PRGS was useful for prognostication.^114^

### Molecular characteristics of appendiceal tumors

Molecular characteristics of appendiceal tumors are not yet well-understood, but rapidly accelerating genomic research suggests that tumor genomics are associated with pathological appearance and that molecular testing has a role in patient stratification.

Some major single mutations in appendiceal tumors have been well-characterized. A recent systematic review of genomic analysis of over a thousand appendiceal tumors confirmed the five most commonly affected genes to be *KRAS, GNAS, TP53, APC*, and *SMAD4*, with relative frequencies distinct from those of colorectal cancer, and also clear differences between appendiceal tumors with goblet cell vs. mucinous histology.^115^ *GNAS* mutation has consistently been associated with low-grade mucinous tumors and *TP53* associated with high-grade tumors and worse overall prognosis..^116^ Rapidly advancing genomic analysis techniques provide a host of new clues about tumor behavior – such as the poor prognosis of tumors overexpressing genes associated with stem-cell-like behavior; newly suspected tumor driver mutations in *HERC2* and *TGFBR2*; and the negative prognostic impact of mutations in the *CDKN2A* and *mTOR* genes.^117,118^

Increasingly, focus has shifted to identifying tumor molecular profiles or mutation clusters that provide insight into tumorigenesis, survival, and treatment response.^118,119^ One key study generated four molecular subtypes of appendiceal adenocarcinoma, of which *RAS*-mutation predominant tumors had the most favorable survival and the greatest likelihood of responding to chemotherapy, while *TP53*-predominant tumors were associated with greater genomic complexity and inferior outcomes.^120^ Molecular subtype was independently associated with survival, even beyond histologic type and grade.^120^ Another novel study grouped tumors as oncogene-enriched, immune-enriched, or intermediate, and demonstrated the lowest-risk behavior in immune-enriched tumors, suggesting a major component of immune dysregulation in appendiceal tumorigenesis.^117^ Finally, machine-learning approaches have allowed for more objective cluster identification. Garland-Kledzik and colleagues identified 5 novel mutation subtypes that suggested tumorigenesis patterns, including one with mutations associated primarily with epigenomic reprogramming, and one without any of the currently known common mutations, suggesting an as-yet unidentified driver of tumorigenesis.^121^

### Pathology-Based Treatment Guidelines

Primary and peritoneal grades and types of appendix tumors have been arranged into a grid of all possible combinations in Table 4, with the corresponding treatment recommendations for surgical resection of the primary, cytoreduction, and systemic chemotherapy. Non-mucinous and GCA are addressed together, as in the consensus pathway. Neuroendocrine tumors are not addressed in this guideline.

**Table 4.**
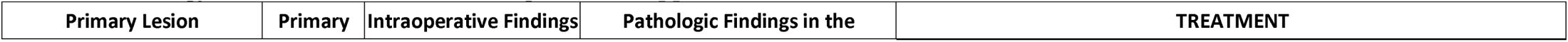
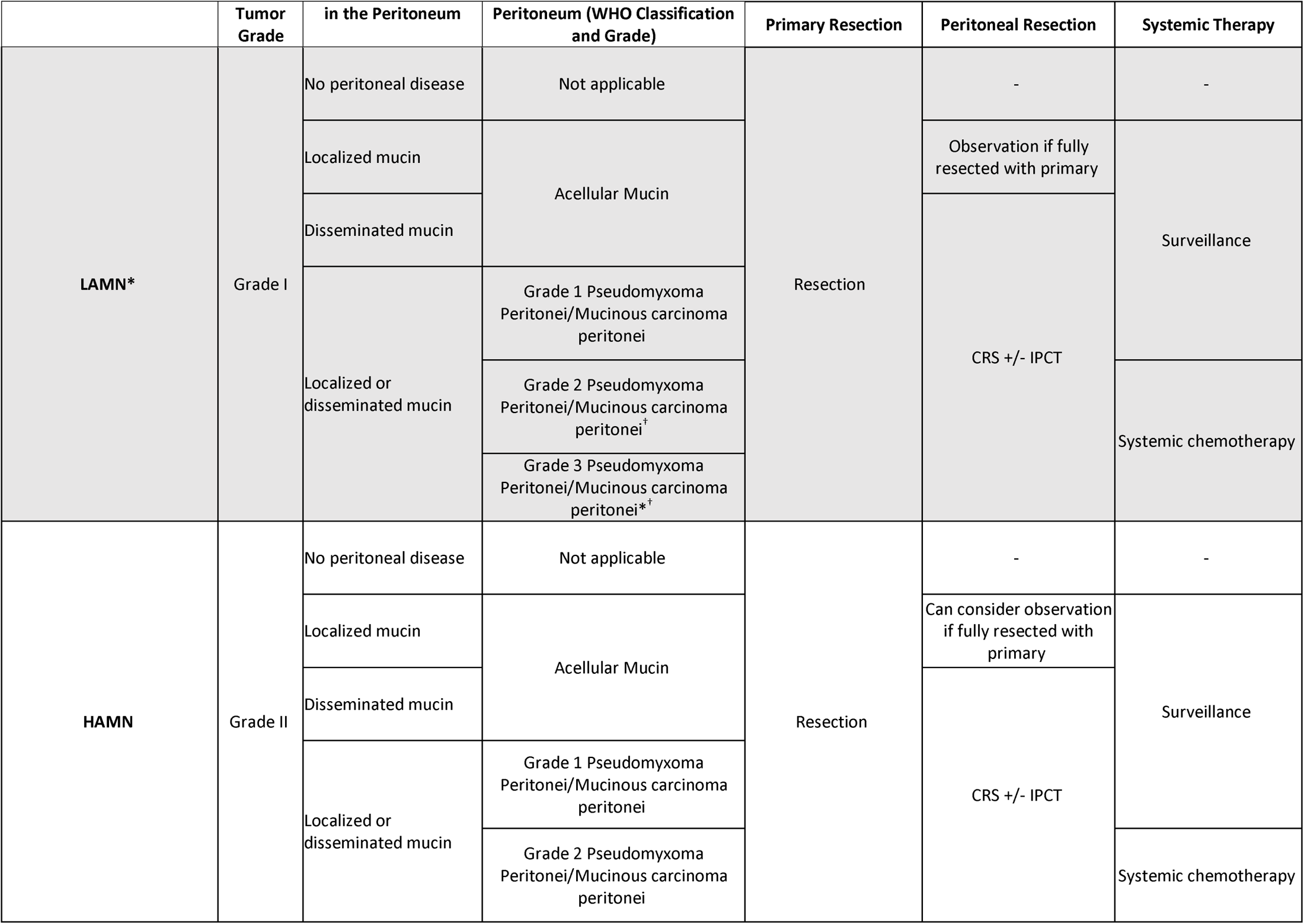
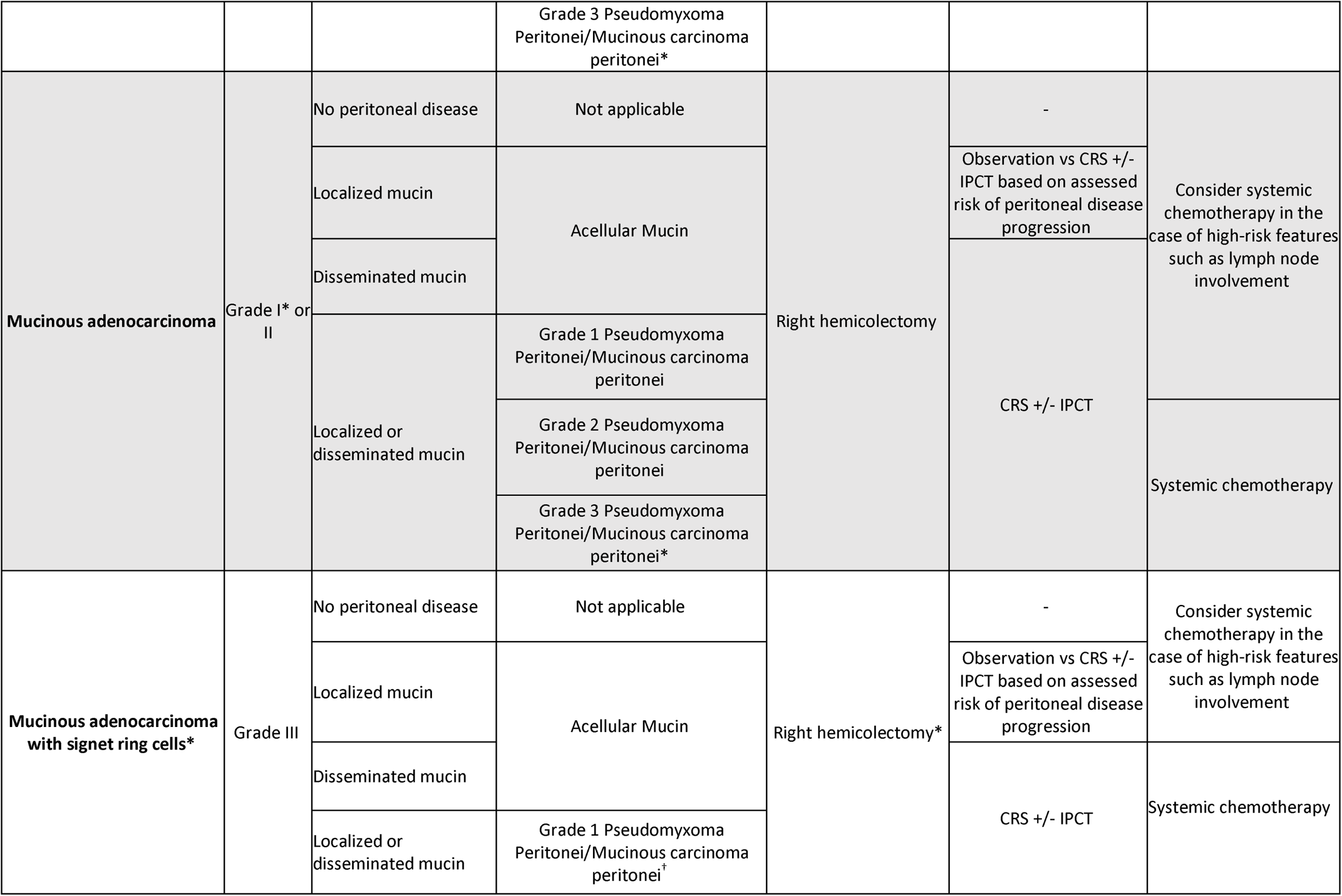
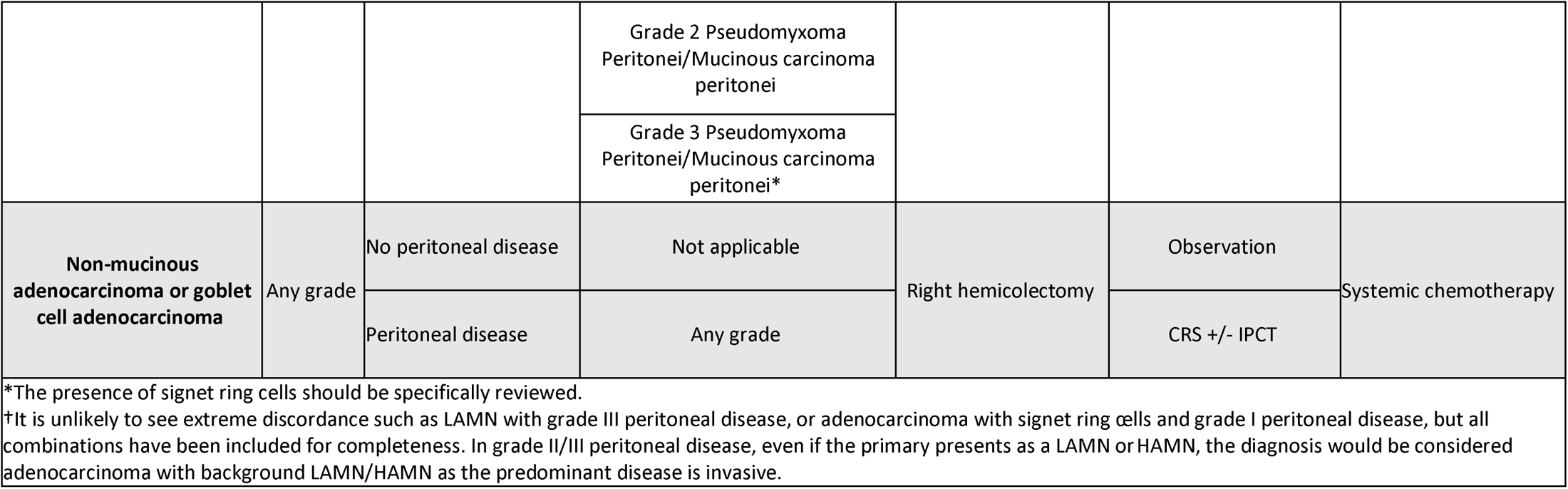
Pathology-Based Guidelines for Management of Appendiceal Tumors.

All three consensus pathways are summarized in this table; the recommendations will not be recapitulated in full here, but notably, for AMNs, the primary should be resected to negative margins in the least invasive safe fashion. For LAMN, extra-appendiceal acellular mucin that is localized to the meso-appendiceal fold and peri-appendiceal recesses can be managed by complete resection alone. If extra-appendiceal mucin is cellular or more extensively disseminated, cytoreduction with or without intraperitoneal chemotherapy should be offered to surgical candidates. ^25,27,122–128^ Recommendations for HAMN differ from LAMN only in that cytoreduction may be considered in localized extra-appendiceal acellular mucin, given higher rates of progression and recurrence.^21^ Cytotoxic chemotherapy should be considered only for grade II or III peritoneal disease, as no benefit has been demonstrated for grade I disease. ^57,129–135^ Emerging evidence suggests atezolizumab/bevacizumab combination therapy may be considered for unresectable disease of any grade, as discussed in part 1.^57^

For mucinous adenocarcinoma, right hemicolectomy should be performed, with the exception of well-differentiated adenocarcinoma confined to the appendix, as described in part 1.^15,136–138^ Cytoreduction with or without intraperitoneal chemotherapy may be considered for localized acellular mucin, based on the patient’s assessed risk of peritoneal disease progression; it should be pursued for disseminated acellular mucin and all cellular mucin.^25,27,122–128^ Systemic chemotherapy should be considered if the primary adenocarcinoma has high-risk features as described in the part 1 guideline, including lymph node involvement or signet ring cells, or for grade II or III peritoneal disease. ^,20,22,45,70,129–135,139–143^

For GCA or non-mucinous adenocarcinoma, right hemicolectomy should be performed. If any peritoneal disease is present, cytoreduction should be performed. Systemic chemotherapy should be administered prior to cytoreduction if possible.^129–135,142^

## APPENDICEAL TUMORS WITH PERITONEAL DISEASE

### Consensus Results and Updates

The pathway’s nine final main blocks are summarized below (Figure 1). The most important update from 2018 is stratification of recommendations by pathologic grade of peritoneal disease for mucinous disease. All non-mucinous disease is grouped together. An overarching change in treatment recommendations is the preference in most instances for a period of SCT prior to cytoreduction. As with localized tumors, initial workup recommendations are more comprehensive, and surveillance recommendations are unified and updated. A tenth block describing novel therapies was proposed for the Delphi 1 consensus voting but was discarded, as many of the 77% who approved of the content expressed a preference to exclude novel therapies from a guideline algorithm. Agreement tables can be reviewed in Table 5.

**Figure 1.**
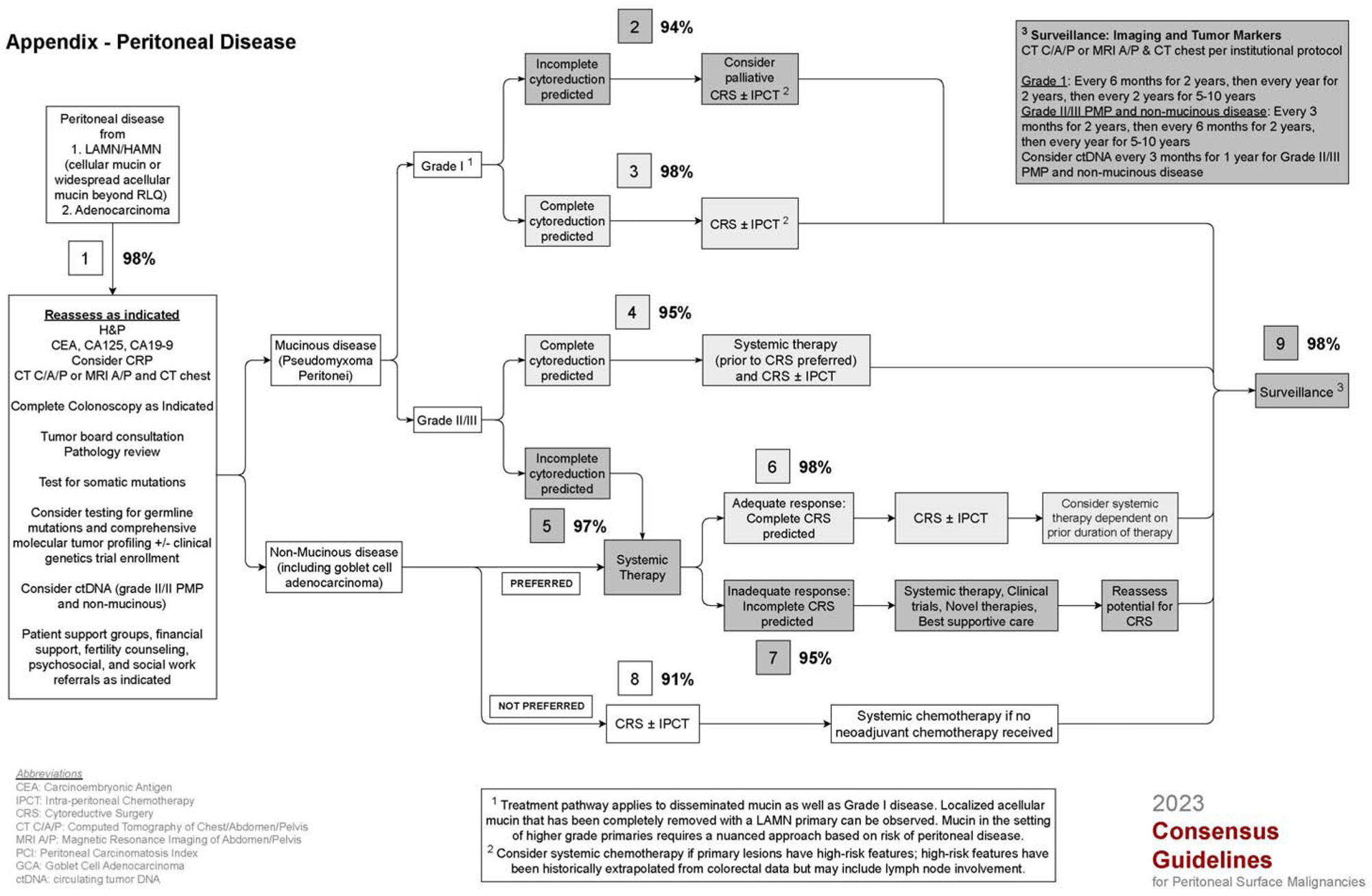
Appendiceal Tumors with Peritoneal Disease Pathway

**Table 5.**
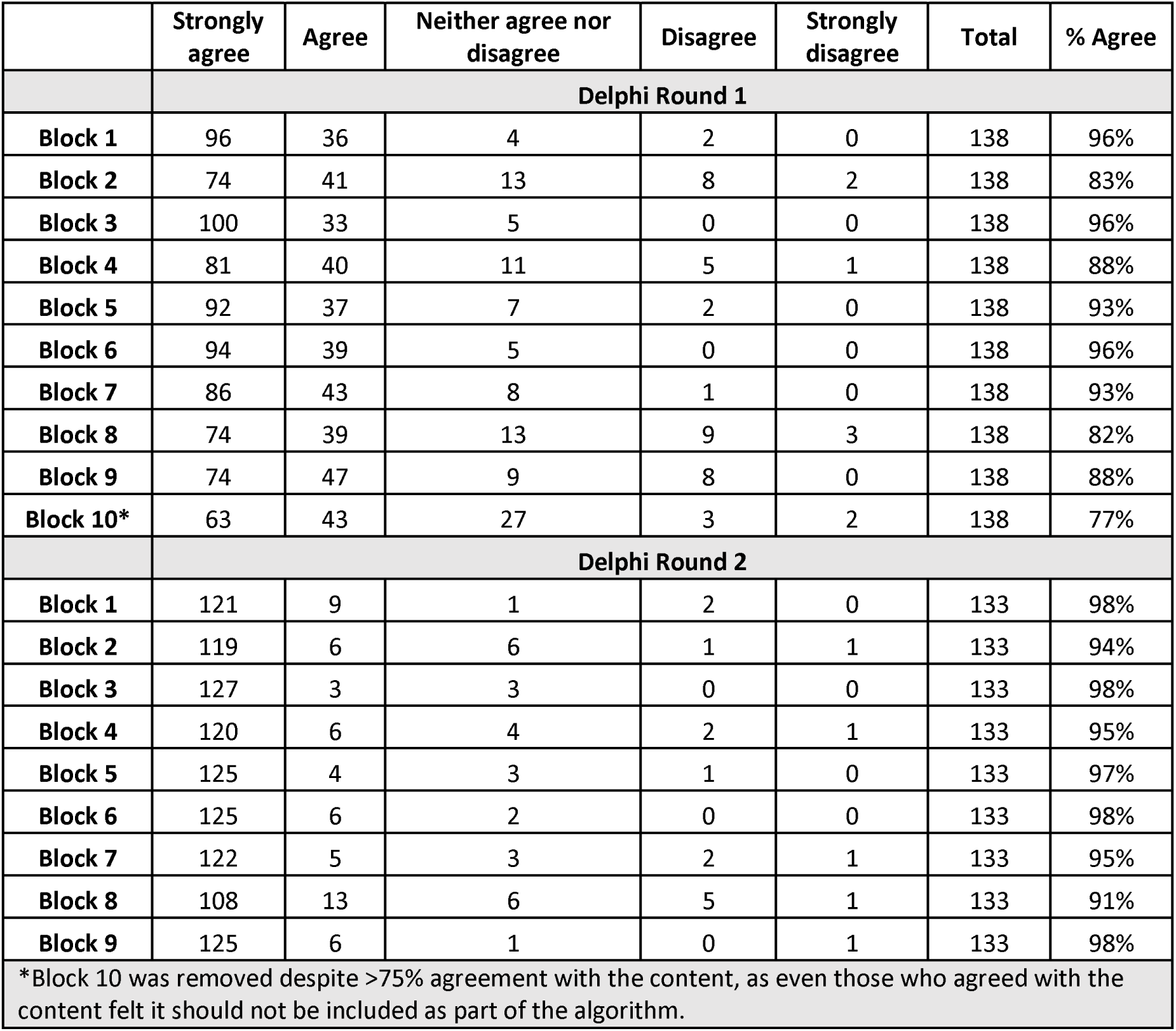
Appendiceal Tumors with Peritoneal Involvement Delphi 1 and 2 Agreement (% agreement includes agree and strongly agree)

#### Block 1

Peritoneal disease of appendiceal origin may present at initial diagnosis, or as progression or recurrence of previously diagnosed disease. If noted intraoperatively, biopsies should be taken at that time; otherwise, laparoscopy should be considered to obtain a tissue diagnosis.

Workup should include either an initial complete (if index diagnosis) or appropriately updated full history and physical including family and personal cancer history and cancer risk factors, tumor markers including CEA, CA-125, and CA 19-9 in addition to consideration of CRP, and completion chest and abdominopelvic cross-sectional imaging.^144–148^ A recent study of over 1300 patients with appendiceal adenocarcinoma demonstrated an association between each of CEA, CA125, and CA19-9 with overall survival, further supporting their inclusion in the workup.^149^ Colonoscopy should be recent or updated.^150,151^ Genomic testing for advanced cancers may include somatic variant profiling with consideration for further germline variant evaluation.^152,153^ For non-low-grade disease, circulating tumor DNA (ctDNA) testing should be considered at baseline for future surveillance purposes.^154^

As with all appendix tumors, patients should be discussed in tumor board and pathology reviewed by an expert pathologist. Patients should be evaluated for additional support needs, including patient support groups, social work, financial support, psychosocial support, and fertility counseling.

**% Agreement**: 96% in first round, 98% in second

#### Block 2

Blocks 2 and 3 address grade I PMP and disseminated acellular mucin associated with any primary other than grade III adenocarcinoma. A plan of care must be nuanced, based on the risk of peritoneal disease suggested by pathologic features and the patient’s surgical fitness and risk profile. In block 2, palliative or temporizing cytoreduction and IPCT should be considered even if incomplete cytoreduction is predicted.

Cytoreduction should include right hemicolectomy when the primary is adenocarcinoma. ^15,136–138^ Systemic chemotherapy has generally not been shown to improve outcomes in grade I disease. However, if the primary tumor has high-risk features such as lymph node involvement or high-grade or signet ring cell histology, SCT is indicated as described in part 1. If the primary is identified to be grade III prior to cytoreduction, consensus recommendation is for preoperative chemotherapy, as in block 4.^20,22,45,70,139–143^

There is not adequate data nor consensus at this time to unilaterally recommend local resection only, more invasive CRS ± IPCT, or SCT for the treatment of limited acellular mucin in the right lower quadrant when the primary disease is adenocarcinoma; however, it may be reasonable to consider observation if fully resected in the absence of high-risk tumor features.

**% Agreement**: 83% in first round, 94% in second

#### Block 3

In block 3, for grade I disease with complete cytoreduction predicted, definitive cytoreduction and intraperitoneal chemotherapy should be performed, including right hemicolectomy for adenocarcinoma. ^15,25,27,122–128,136–138^ Systemic chemotherapy is indicated if the primary tumor has high-risk or high-grade features. ^20,22,45,70,139–143^

**% Agreement**: 96% in first round, 98% in second

#### Block 4

Block 4 addresses grade II/III peritoneal mucinous disease or any PMP with a grade III primary. If complete cytoreduction is predicted, then both CRS ± IPCT and SCT should be carried out.^129–135^ Upfront SCT is preferred by consensus to assess disease response, followed by planned complete cytoreduction, although this is not universal.

**% Agreement**: 88% in first round, 95% in second

#### Block 5

Block 5 addresses grade II/III peritoneal mucinous disease with incomplete cytoreduction predicted, and non-mucinous peritoneal disease. In these cases, chemotherapy should be performed upfront and response assessed prior to further surgical planning. ^129–135,142^

**% Agreement**: 93% in first round, 97% in second

#### Block 6

After patients with peritoneal disease undergo a course of systemic chemotherapy, they should be re-assessed for response. If predicted incomplete cytoreduction converts to predicted complete cytoreduction, or complete cytoreduction remains feasible, then CRS ± IPCT should be pursued. ^129–135,142^ Total duration of chemotherapy must ultimately be determined by a medical oncologist with subject matter expertise but is in most cases recommended for a duration of 6 months, and if not completed preoperatively, should be completed postoperatively.^5,129,155^

**% Agreement**: 96% in first round, 98% in second

#### Block 7

After a course of chemotherapy, if incomplete cytoreduction is persistently predicted or disease has progressed substantially, patients with higher-grade mucinous or any non-mucinous peritoneal pathology should not be offered CRS ± IPCT as definitive therapy. Survival at three years after incomplete cytoreduction for high-grade malignancy is as low as 9%, and it is the consensus opinion that this is unlikely to justify the surgical risks for most patients.^156^ Depending on the initial chemotherapy course and characteristics of disease progression, patients should be referred for further SCT, novel and/or clinical trial therapies, and/or best supportive care. As therapy progresses, patients should be evaluated at regular surveillance intervals for progression or potential for cytoreduction.

Systematic review does indicate that tumor debulking in appropriate surgical patients, be it destination therapy or a bridge to further surgery or intraperitoneal chemotherapy, may have a survival or symptom control benefit. The risks and benefits should be carefully considered on an individualized basis, and patients counseled that this is not a definitive curative therapy if it is offered.

**% Agreement**: 93% in first round, 95% in second

#### Block 8

Upfront CRS ± IPCT is not preferred in non-mucinous peritoneal disease. However, if a patient undergoes upfront surgery, they should receive a full course of SCT postoperatively.

**% Agreement**: 82% in first round, 91% in second

#### Block 9

After the conclusion of blocks 3, 4, 6, 7, or 8, patients should proceed to surveillance protocols. Surveillance, as in every other pathway, includes cross-sectional imaging of the chest, abdomen, and pelvis at regular intervals by either MRI or CT; tumor markers (CEA and any other markers that have been elevated during the disease course); and an updated history and physical.

For grade I peritoneal disease, imaging should be every 6 months for two years, then every year for 2 years, then every 2 years for 5-10 years. For all others, imaging should be every 3 months for two years, every 6 months for two years, and then every year for 5-10 years. This generally follows the US HIPEC collaborative, although by expert consensus it is recommended to have enhanced intensity of surveillance for the higher risk grade II/III disease in the initial post-interventional phase.^157^

Consider monitoring circulating tumor DNA every 3 months for 1 year for grade II/III and non-mucinous disease. There is not yet strong evidence for its use in grade I disease.

**% Agreement**: 88% in first round, 98% in second

## DISCUSSION

A chief benefit of this update is unification of recommendations across a multidisciplinary consensus group and single pathologic grading system, with guidance on how to relate current recommendations to previous classification schemes. Major updates are the new preferential recommendations for timing of chemotherapy and cytoreduction, and unified surveillance recommendations.

As in part 1, limitations of the consensus include the observational nature of the relevant body of evidence. The role of intraperitoneal chemotherapy remains controversial among consensus members and thus we provide no universal recommendation, but several studies in appendix tumors suggest benefit. The increased diversity in expertise represented in this consensus group is a major strength.

## CONCLUSION

Herein is reported an updated Delphi consensus of management guidelines concerning appendiceal tumors with peritoneal involvement. Importantly, this consensus group contained experts across multiple disciplines relevant to cancer care and patient advocates. Cytoreduction remains the bedrock of up-front definitive treatment in low-grade peritoneal disease. Individuals with high-grade disease should first undergo systemic therapy, and if peritoneal disease remains or becomes resectable, should be evaluated for cytoreduction. Supportive multidisciplinary therapies and palliative surgery should be considered whenever they offer a quality-of-life advantage. Key takeaways are highlighted in Table 6.

**Table 6.** Key takeaways.

|  |
| --- |
| The WHO 2019 (5<sup>th</sup> edition) classification system should be used for appendiceal tumors. |
| The crosstalk between this classification system and other relevant systems that may be encountered is presented (Table 3). |
| Management of both localized and advanced appendiceal tumors depends upon tumor histology and grade, in addition to stage. Recommendations are related in the context of specific tumor pathology (Table 4). |
| For appendiceal tumors with peritoneal involvement, we recommend consideration of CRS with or without IPCT for patients with resectable disease, and systemic chemotherapy for patients with histology other than low-grade, in the sequence represented in the Figure 1 care pathway. |
| The optimal management of both initially unresectable and recurrent peritoneal disease remains uncertain; results of a systematic review suggest components of a multidisciplinary approach that may have benefit in carefully selected patients, but bias cannot be ruled out and additional study is needed. |

## Data Availability

All data produced in the present study are available upon reasonable request to the authors

