## Supplemental Tables 1-6, Figures 1-3 for "Consensus guideline for the management of patients with appendiceal tumors: Part 2: Appendiceal tumors with peritoneal involvement"

### Supplemental Material

#### Supplemental Table 1. Search Strategy for Key Question 2

PubMed; restrictions: any date - 2023/06/15, performed on humans

| **Search line** | Search term |
| --- | --- |
| **1** | Appendi*[tw] AND (cancer*[tw] OR adenocarcinoma*[tw] OR carcinoma*[tw] OR neoplasm*[tw]) OR "goblet cell"[tw] OR (Appendiceal neoplasms[MeSH Major Topic]) OR pseudomyxoma[tw] OR "pseudomyxoma peritonei"[MeSH Major Topic] |
| **2** | Unresectable[tw] OR palliat*[tw] OR Replase* OR recur*[tw] OR Extensive*[tw] OR advanced*[tw] OR progress*[tw] OR "high burden"[tiab:~5] OR "high PCI"[tiab:~5] OR diffuse*[tw] OR "Palliative Medicine"[Mesh] OR "Palliative Care"[Mesh] |
| **3** | Evacuat*[tw] OR paracent*[tw] OR cathet*[tw] OR debulk*[tw] OR ("surgery peritoneal"[tiab:~5]) OR ("surgery peritoneum"[tiab:~5]) OR ("resection peritoneal"[tiab:~5]) OR ("resection peritoneum"[tiab:~5]) OR incomplete cytoreduc*[tw] OR incomplete CRS[tw] OR intrapertioneal chemotherapy[tw] OR HIPEC[tw] OR Neoadjuvant[tw] OR Adjuvant[tw] OR Perioperative[tw] OR Chemotherapy OR Immunotherapy[tw] OR Targeted therapy[tw] OR Systemic therapy[tw] OR Drug Therapy[MeSH Major Topic] OR Antineoplastic agents[MeSH Major Topic] OR Drug therapy, Combination[MeSH Major Topic] OR chemotherapy, adjuvant[MeSH Major Topic] OR neoadjuvant therapy[MeSH Major Topic] |
| **4** | #1 AND #2 AND #3 |

#### Supplemental Table 2. Search Strategy for Key Question 3

PubMed; restrictions: any date - 2023/06/15, performed on humans

| **Search line** | Search term |
| --- | --- |
| **1** | Appendi*[tw] AND (cancer*[tw] OR adenocarcinoma*[tw] OR carcinoma*[tw] OR neoplasm*[tw]) OR "goblet cell"[tw] OR (Appendiceal neoplasms[MeSH Major Topic]) OR pseudomyxoma[tw] OR "pseudomyxoma peritonei"[MeSH Major Topic] |
| **2** | (Colon cancer OR Rectal cancer OR Colorectal cancer OR Colon neoplasm* OR Rectal neoplasm* OR Colorectal neoplasm* OR Colon tumor* OR Rectal tumor* OR Colorectal tumor* OR Colon tumour* OR Rectal tumour* OR Colorectal tumour* OR (Colorectal neoplasms[MeSH Major Topic]) OR (Colorectal Surgery[MeSH Major Topic]) OR (Gastrointestinal neoplasms[MeSH Major Topic])) |
| **3** | repeat*[tw] OR iterat*[tw] |
| **4** | Cytoreduct*[tw] OR CRS[tw] OR intraperitoneal chemotherapy[tw] OR HIPEC[tw] OR (Hyperthermic Intraperitoneal Chemotherapy[MeSH Major Topic]) |
| **5** | #1 OR #2 |
| **6** | #3 AND #4 AND #5 |

#### Supplemental Figure 1. Key Question 2 Prisma Flow Diagram

Studies from databases/registers **(n = 1476)**

MEDLINE (n = 1469)

Citation searching (n = 7)

**Identification**

Included studies ongoing **(n = 0)**

Studies awaiting classification **(n = 0)**

Studies included in review **(n = 15)**

Studies excluded **(n = 1370)**

Studies not retrieved **(n = 0)**

Studies assessed for eligibility **(n = 103)**

Studies sought for retrieval **(n = 103)**

Studies screened **(n = 1473)**

Studies excluded **(n = 88)**

Not in English (n = 3)

Wrong outcomes (n = 3)

Wrong study design (n = 25)

Wrong patient population (n = 57)

References removed **(n = 3)**

Duplicates identified by Covidence (n = 3)

**Included**

**Screening**

#### Supplemental Table 3. Inclusion Criteria and Review Materials for KQ2

Reasons for exclusion from any key question review overlap significantly. Primary areas of difference between key questions are the populations, interventions, and outcomes of interest.

| Reason | Criteria | EXclusions |
| --- | --- | --- |
| Language/availability | Available in English | Not able to obtain abstract in English |
| Study design | Original research  Cohort or case-control study, or single-arm or randomized interventional trial | Case series or report  Population overlaps with another study/population and cannot be clearly delineated |
| Population | Individuals with non-recurrent appendix tumors with peritoneal involvement that is assessed to be unresectable | Population does not have mucinous peritoneal disease of appendiceal origin, or peritoneal disease is completely resectable  Population also includes recurrent disease |
| Intervention | Any surgical, procedural, or medical intervention targeting PMP | No description or sub-analysis of receipt of PMP-targeted interventions  Other interventions or diseases are present that would confound analysis (other surgeries, cancers) |
| Comparator | No comparator required for raw survival time  Can compare to any intervention or none/supportive care  If outcomes restricted to comparative metrics, must be compared to another eligible patient group | Comparative metrics only, with comparator group that is ineligible for reasons of disease type or burden, or unclear overlap with treatment group |
| Outcome | Outcomes of interest: overall and progression-free survival, eligibility for and completeness of cytoreduction, and adverse/treatment effects | No description or sub-analysis of outcomes by treatment, disease, or unresectable group (such as outcomes that cannot be extracted from fully resectable disease or mixed treatments) |

#### Supplemental Table 4. Quality Assessment of Included Studies for KQ2

| Quality assessment of studies assessing optimal management approaches for initially unresectable PSM of appendiceal origin | | | | |
| --- | --- | --- | --- | --- |
| Author | **Selection** | **Comparability** | **Outcomes** | **Overall** |
| Sideris 2009^1^ | *** | NA | ** | ***** |
| Smeenk 2007^2^ | *** | NA | *** | ****** |
| Murphy 2007^3^ | **** | Does not control | *** | ******* |
| Farquharson 2008^4^ | *** | NA | *** | ****** |
| Shapiro 2010^5^ | **** | Does not control | *** | ******* |
| Dayal 2013^6^ | *** | NA | *** | ****** |
| Choe 2015^7^ | **** | Does not control | *** | ******* |
| Polanco 2016^8^ | *** | NA | *** | ****** |
| Delhorme 2016^9^ | *** | NA | *** | ****** |
| Berger 2021^10^ | **** | Does not control | *** | ******* |
| Trilling 2021^11^ | *** | NA | ** | ***** |
| Mangieri 2022^12^ | **** | Does not control | *** | ******* |
| Vierra 2022^13^ | *** | NA | ** | ***** |
| Baron 2022^14^ | **** | Does not control | *** | ******* |
| Sugarbaker 2022^15^ | **** | Does not control | *** | ******* |

#### Supplemental Figure 2. Key Question 3 Prisma Flow Diagram

Studies from databases/registers **(n = 193)**

MEDLINE (n = 182)

Citation searching (n = 11)

Included studies ongoing **(n = 0)**

Studies awaiting classification **(n = 0)**

Studies included in review **(n = 9)**

Studies excluded **(n = 155)**

Studies not retrieved **(n = 0)**

Studies assessed for eligibility **(n = 36)**

Studies sought for retrieval **(n = 36)**

Studies screened **(n = 191)**

Studies excluded **(n = 27)**

Wrong outcomes (n = 2)

Low sample size (n = 3)

Wrong patient population (n = 13)

Potential overlap with another study from the same institution (n = 9)

**Identification**

References removed **(n = 2)**

Duplicates identified by Covidence (n = 2)

**Screening**

**Included**

#### Supplemental Table 5. Inclusion Criteria and Review Materials for KQ3

Reasons for exclusion from any key question review overlap significantly. Primary areas of difference between key questions are the populations, interventions, and outcomes of interest.

| Reason | Criteria | EXclusions |
| --- | --- | --- |
| Language/availability | Available in English | Not able to obtain abstract in English |
| Study design | Original research  Cohort or case-control study, or single-arm or randomized interventional trial | Case series or report  Population overlaps with another study/population and cannot be clearly delineated |
| Population | Individuals with recurrent appendix tumors with mucinous peritoneal involvement after previous complete (CC-0 or CC-1 equivalent) cytoreduction | Population does not have mucinous peritoneal disease of appendiceal origin or is index rather than recurrent disease  Population is mixed with previously unresected (progressive) disease |
| Intervention | Repeat cytoreduction with or without intraperitoneal chemotherapy | No description or sub-analysis of intervention group  Other interventions or diseases are present that would confound analysis (other surgeries, cancers) |
| Comparator | No comparator required for raw survival time  Can compare to other common interventions (chemotherapy) or none  If outcomes restricted to comparative metrics, must be compared to another eligible patient group | Comparative metrics only, with comparator group that is ineligible for reasons of disease type or presentation, rare or poorly-defined treatment (novel therapy only, etc), or unclear overlap with treatment group |
| Outcome | Outcomes of interest: overall survival, adverse/treatment effects | No description or sub-analysis of outcomes by treatment, disease, or recurrence group (such as outcomes that cannot be extracted from colorectal cancer or progressive disease) |

#### Supplemental Table 6. Quality Assessment of Included Studies for KQ3

| **Quality assessment of studies assessing efficacy and safety of repeat cytoreduction for recurrent peritoneal surface malignancies of appendiceal origin.** | | | | |
| --- | --- | --- | --- | --- |
| **Author** | **Selection** | **Comparability** | **Outcomes** | **Overall** |
| Ahmadi 2021^16^ | **** | ** | ** | ******** |
| Delhorme 2017^17^ | **** | Does not control | ** | ****** |
| Karpes 2020^18^ | **** | ** | *** | ********* |
| Lopez-Ramirez 2022^19^ | **** | ** | *** | ********* |
| Valenzuela 2022^20^ | **** | Does not control | *** | ******* |
| Kitai 2020^21^ | **** | Does not control | *** | ******* |
| Kong 2021^22^ | **** | Does not control | * | ***** |
| Powers 2020^23^ | **** | Does not control | * | ***** |
| Yan 2007^24^ | **** | Does not control | *** | ******* |

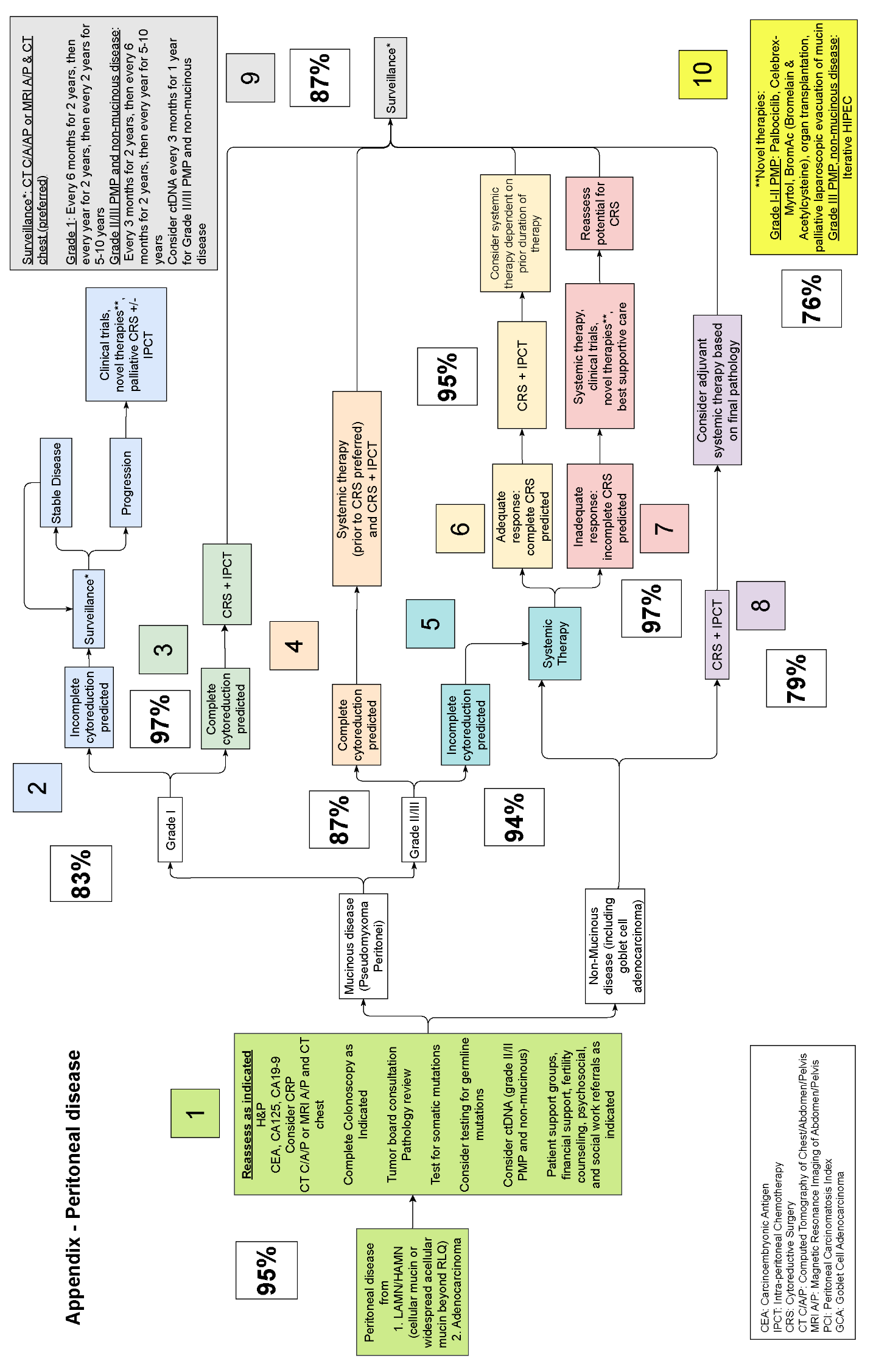

#### Supplemental Figure 3. Round 1 Appendiceal Peritoneal Disease Pathway
